# 2022 Sudan Ebolavirus Outbreak in Uganda: Modelling Case Burden and Outbreak Duration

**DOI:** 10.1101/2024.10.11.24314870

**Authors:** Donal Bisanzio, Henry Kyobe Bosa, Barnabas Bakamutumaho, Carolyne Nasimiyu, Diana Atwine, Daniel Kyabayinze, Charles Olaro, Robert F. Breiman, M. Kariuki Njenga, Henry Mwebesa, Jane Ruth Aceng, Richard Reithinger

## Abstract

In September 2022, an outbreak of Sudan virus (SUDV) was confirmed in Uganda. Following the first case report, we developed an individual based modelling platform (IBM-SUDV) to estimate the burden of cases and deaths, as well as the duration of the unfolding SUDV outbreak, using different scenarios. Modelled projections were within the range of cases and deaths ultimately observed.

## INTRODUCTION

Ebola virus disease (EVD) is a severe, often fatal illness affecting humans and other primates. It is caused by is caused by a group of viruses, known as orthoebolaviruses (family *Filoviridae*).[1] In the past four decades there have been 36 Ebola virus disease (EVD) outbreaks across 11 countries, resulting in >15,000 deaths.[2] With a case fatality rate of more than 65%, EVD is among the most lethal viral haemorrhagic fevers. Consequently, when outbreaks occur, affected countries’ Ministries of Health (MOHs) and regional public health emergency responders are put on high alert.

On September 20, 2022, an outbreak of *Orthoebolavirus sudanense* (Sudan virus [SUDV]) was confirmed by the Uganda Ministry of Health (MOH) in southcentral Mubende District, Uganda.[3, 4] Following this first case report, the virus rapidly spread to 8 nearby districts over the following weeks. In October, concerns within the MOH and the international community about the potential magnitude of the outbreak accelerated when a treatment-seeking infected individual travelled to the highly populated capital city, Kampala; many new cases were linked to this patient,[5] who eventually died. Because no effective treatment or vaccine existed against SUDV,[6] the MOH’s response to mitigate the outbreak relied on non-pharmaceutical interventions (NPIs), including: aggressive case isolation and contact tracing; safe burials; hygiene promotion; social and behavior change; and lockdowns. These NPIs were applied based on successful experiences from previous EVD outbreaks in sub-Saharan Africa, and built upon the ongoing COVID-19 pandemic response infrastructure.[7]

The aim of the study here was to develop an individual based model (IBM-SUDV) to (i) predict the burden of cases and deaths, as well as (ii) the duration of the unfolding SUDV outbreak in Uganda.

Additionally, we evaluated the possible effect of NPIs on SUDV outbreak dynamics.

## THE STUDY

### Model overview

The IBM-SUDV followed a novel framework recently used to estimate disease burden for COVID-19,[8] mpox,[9] and Ebola [10]. The framework accounts for important factors beyond those included in previously published IBMs for Ebola,[11] including the geographical distribution of the Ugandan population, human movement, heterogeneity of human contact with country-specific demographic characteristics, interaction within households and in the community, interaction among age groups, and the structure of the health system workforce.

In this study, the IBM-SUDV applied a contact network representing human interactions at local and regional levels within the Ugandan population. The network was built using available demographic data (e.g., population age structure, gender ratio, household size) and accounted for heterogeneity in interactions among age groups. The IBM-SUDV included uptake and impact of NPIs simulating the response to the 2022 Ebola outbreak in Uganda, such as contact tracing, isolation, safe burial, and use of personal protective equipment (PPE). Timing of intervention deployment and heterogeneity of response across Uganda were modeled to estimate the impact of the outbreak, specifically with regards to case burden, deaths, and duration of the outbreak.

SUDV transmission in the IBM-SUDV was modeled using the classical SEIR compartmental model structure: Susceptible (S) → Exposed (E) → Infectious (I) → Recovered (R).[8–10] Infectious individuals were at risk of dying from SUDV, and individuals who died remained infectious until they were buried. The transition from one status to another was a function of pathogen characteristics (e.g., probability of effective transmission per close contact, incubation period, infectious period, and fatality rate) and interaction among individuals (only for S to I). For all individuals, parameter values for treatment-seeking behavior, hospitalization, fatality, and burial for the Uganda were determined using either published or assumed data—this represented the Baseline Scenario.

Key IDM-SUDV parameters are listed in **Tables 1–3**. A full description of the IBM-SUDV elements, data, and assumptions is provided in the **Supplementary Appendix**.

**Table 1.**
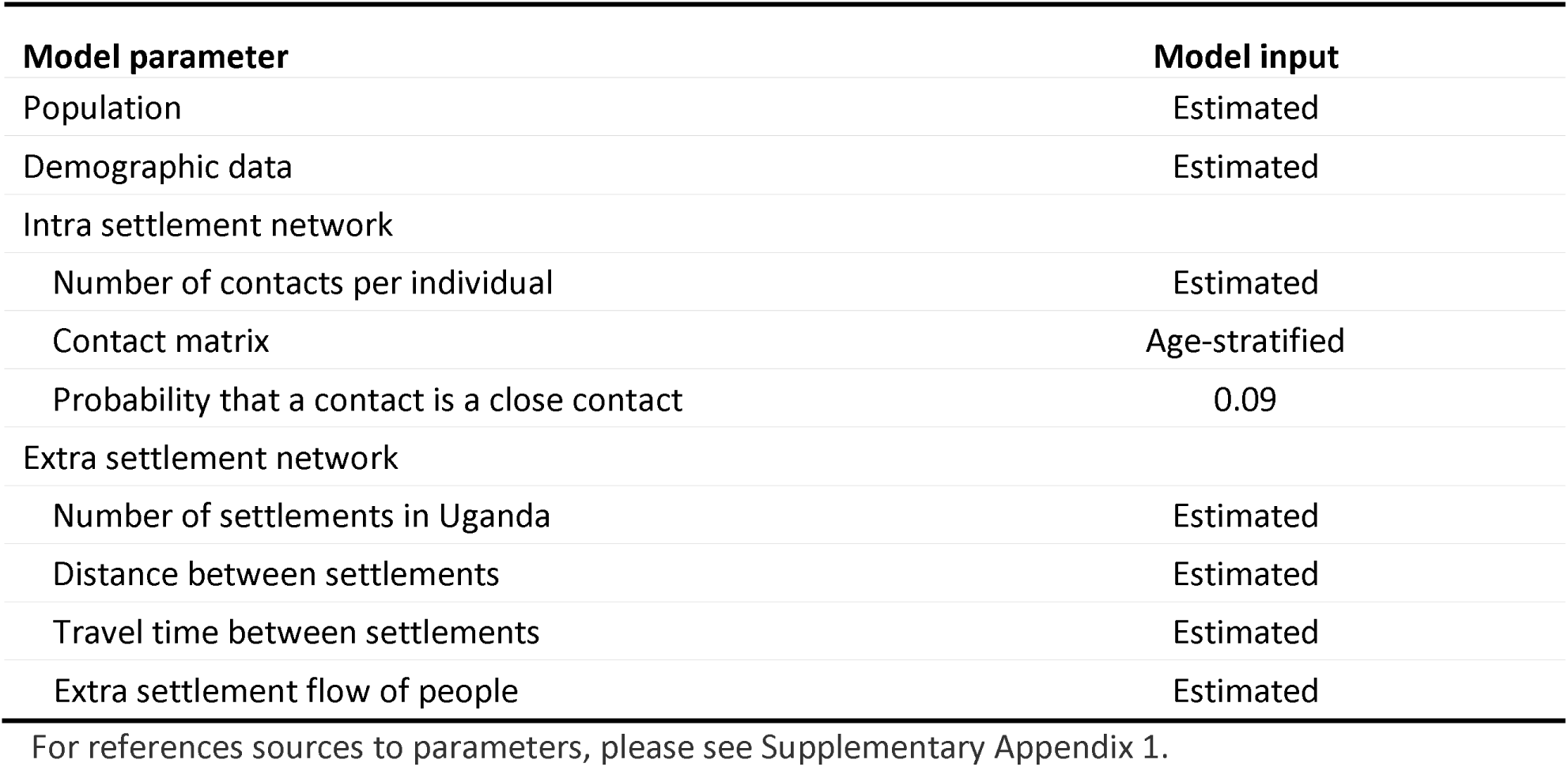
IBM Population Network Parameters and Data Sources.

| **Model parameter** | **Model input** |
| --- | --- |
| Population | Estimated |
| Demographic data | Estimated |
| Intra settlement network |  |
| Number of contacts per individual | Estimated |
| Contact matrix | Age-stratified |
| Probability that a contact is a close contact | 0.09 |
| Extra settlement network |  |
| Number of settlements in Uganda | Estimated |
| Distance between settlements | Estimated |
| Travel time between settlements | Estimated |
| Extra settlement flow of people | Estimated |

For references sources to parameters, please see Supplementary Appendix 1.

**Table 2.** SUDV Transmission Parameters and Data Sources Model parameter Model input.

| **Model parameter** | **Model input** |
| --- | --- |
| Probability of effective transmission per close contact | 0.09–0.12<br>Uniform distribution |
| Incubation period (days to symptom onset) | Mean: 12 days<br>SD: 4.3 days<br>Lognormal distribution |
| Infectious period | 2–21 days<br>Uniform distribution |
| Percentage of asymptomatic cases | 20% |
| Time from symptom onset to hospitalization | 5 days |
| Days in hospital | 10–15 days<br>Uniform distribution |
| Case fatality rate | 39% |
| Time from symptom onset to death | 10 days |
| Days infectious before burial | 7 days |

SD = standard deviation. For references sources to parameters, please see Supplementary Appendix 1.

**Table 3.** Parameters of SUDV Interventions.

| **Model parameter** | **Model input** |
| --- | --- |
| Reduction in transmission probability due to use of PPE at health facilities during the post-intervention period | 70% |
| Probability of seeking treatment at a medical facility in the pre-intervention period | 0.6 |
| Probability of seeking treatment at a medical facility in the post-intervention period | 0.8 |
| Probability of hospitalization among individuals seeking treatment in the pre-intervention period | 0.60 |
| Probability of hospitalization among individuals seeking treatment in the post-intervention period | 0.90 |
| Among deaths in the hospital, probability of having a safe burial | 1.0 |
| Among deaths at home, probability of having a safe burial in the pre-intervention period | 0.1 |
| Among deaths at home, probability of having a safe burial in the post-intervention period | 0.95 |
| Percentage of contacts (first ring) of infected individuals located by contact tracing team | Beta distribution with mean 70% |

PPE = personal protection equipment. For references sources to parameters, please see Supplementary Appendix 1.

### Scenarios

The IBD-SUDV Baseline Model was compared to two hypothetical scenarios:

1. A “Delayed Outbreak Response Scenario”, which assumed a 5-month delay in the response due to timing required to have NPIs in place (i.e., contact tracing, isolation, PPE).
2. An “Out-of-Control Outbreak Scenario”, which assumed a 5-months delay in the response due to timing to have NPIs in place (i.e., contact tracing, isolation, PPE), as well as a 50% contact tracing and isolation rate (i.e., similar to what was observed in the early phase of the 2014–2016 West Africa Ebola outbreak).

Again, elements, data and assumptions for the two scenarios are provided in the **Supplementary Appendix**.

### Simulations and Analyses

For each scenario, 1,000 simulations were completed with a time horizon of 150 weeks each. Simulation results were used to estimate the median number of cases, hospitalizations, and deaths as well as the median duration of the epidemic for each scenario analyzed. The epidemic duration was measured from the occurrence of the first case to the time at which zero cases were observed 42 days after the last infection event. A 95 % credible interval (CrI) was calculated for each outcome using the adjusted bootstrap percentile approach.[11]

## RESULTS

Figure 1 shows the epidemic curve for the baseline and tested scenarios. With NPIs implemented to reduce SUDV transmission, the IBM-SUDV Baseline Scenario estimated a mean number of cases and deaths equal to 193 (95 % CI, 131–277) and 81 deaths (95 % CI, 55–124), respectively. The median duration of the epidemic was 22 weeks (95 % CI, 18–25) (Figure 1A).

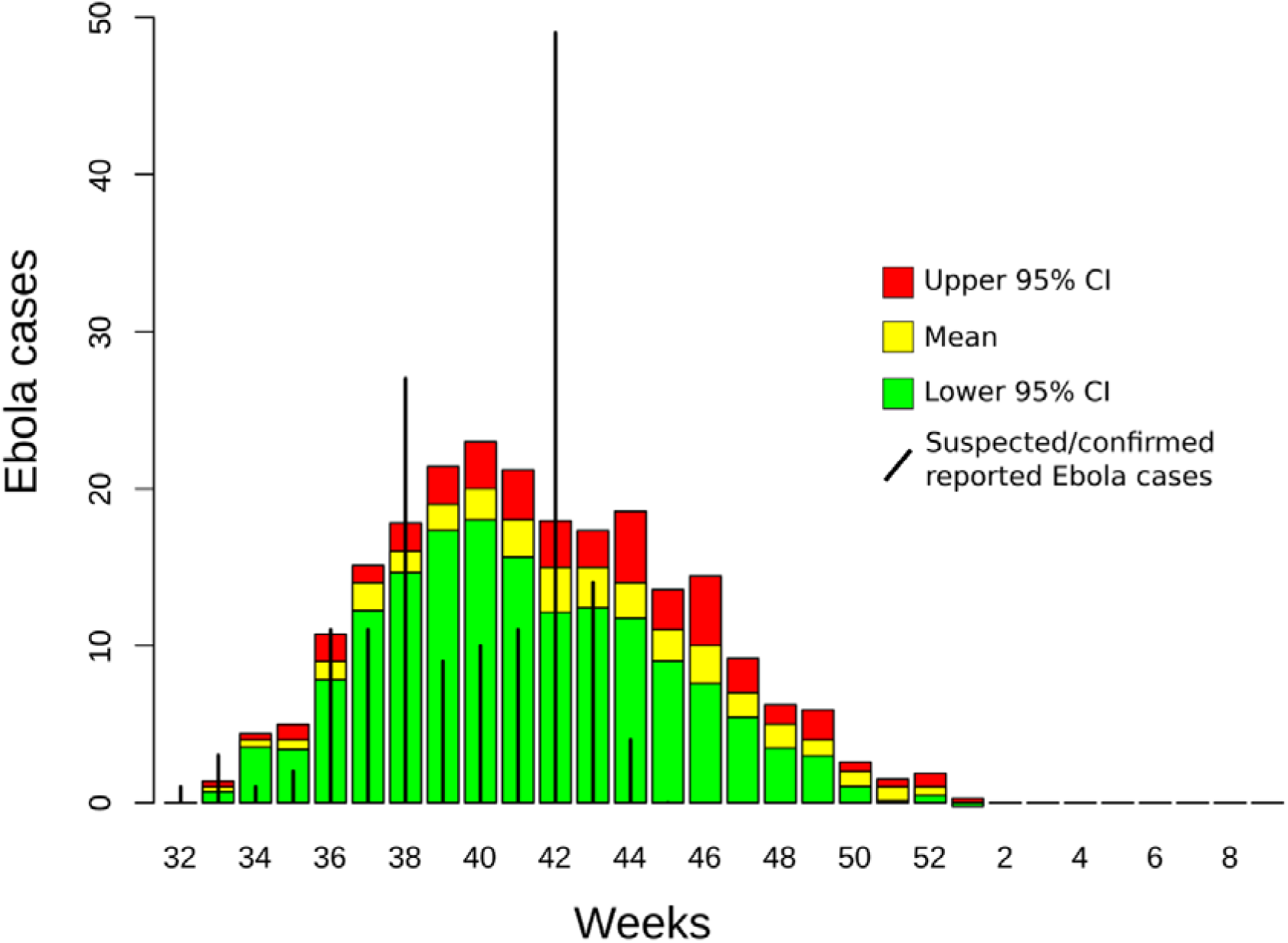
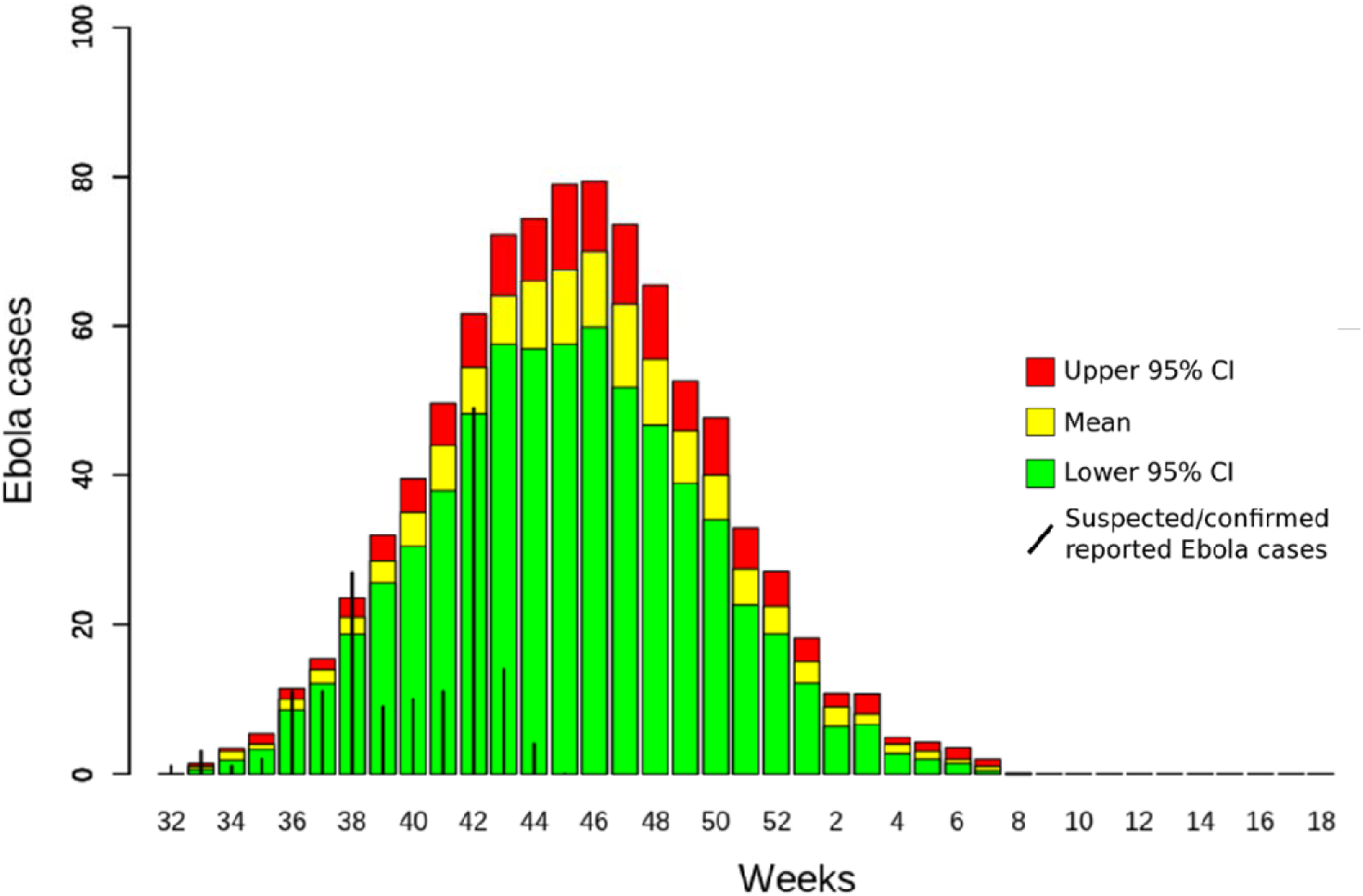
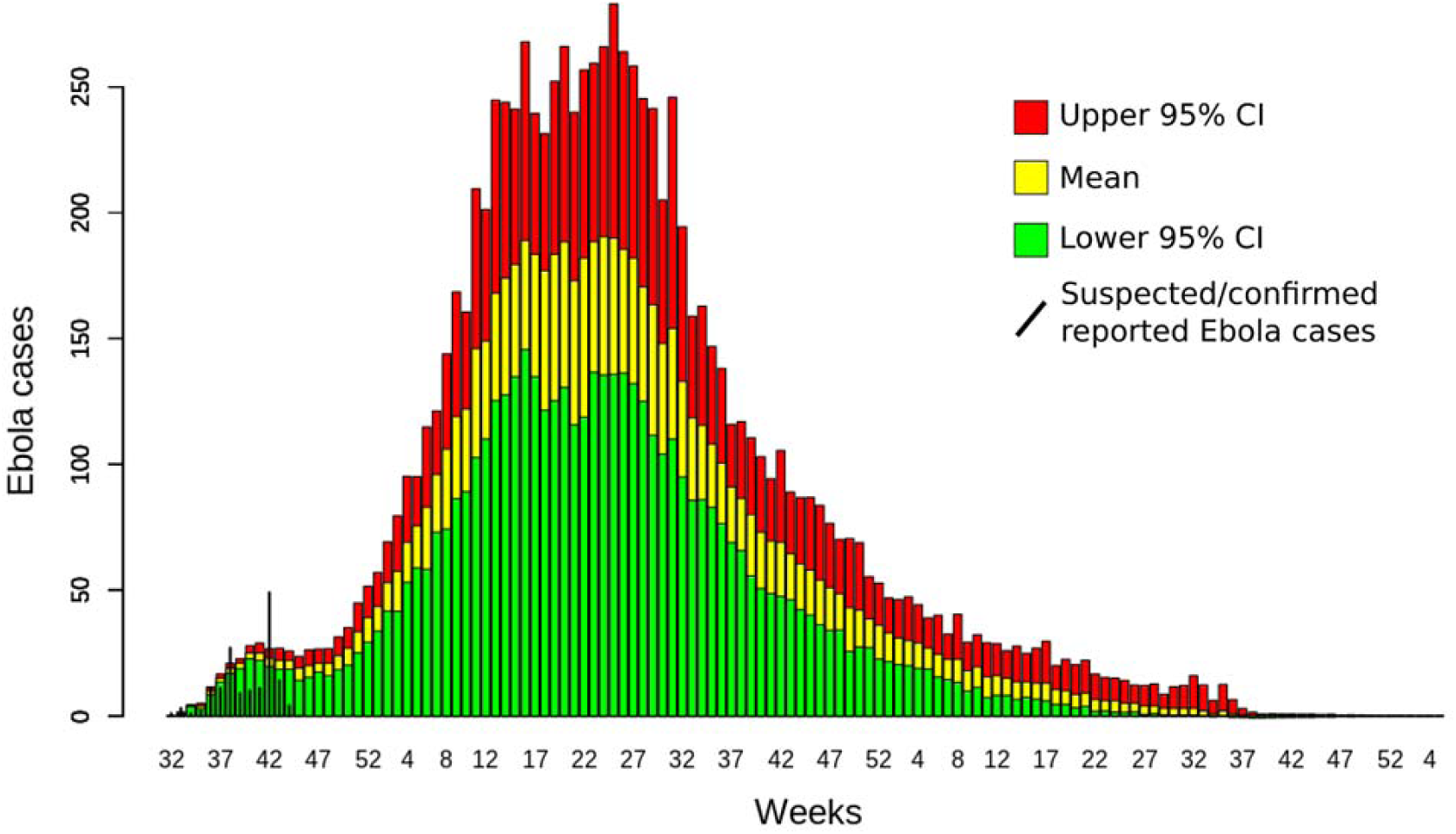

We estimated that the Delayed Outbreak Response Scenario would result in 778 (95 % CI, 665–901) cases and 303 (95 % CI, 259–351) deaths, as well as extending the median duration of the epidemic to 24 weeks (95% CI 20–28) (Figure 1B). An Out-of-Control Outbreak Scenario would result in 13,537 (95 % CI, 9,376– 19,919) cases and 5,279 (95 % CI, 3,656–7,768) deaths, and a median epidemic duration of 24 months (95% CI 22–27) (Figure 1C).

The IBM-SUDV’s modelled projections were completed on November 1, 2022, and shared with the Ugandan MOH on November 11, 2022.

## DISCUSSION

Prior to the 2022 outbreak, Uganda had reported 6 EVD outbreaks, the first one of which had occurred in 2000.[2] Of reported outbreaks, 4 were due to SUDV (2000, 2011, and 2012 [n = 2]), one was due to *O. bundibugyoense* (2007), and one was due to *O. zairense* (2019). The median number of cases and deaths of these outbreaks was 149 (range = 1–596) and 37 (range = 1–275), respectively; in total, the outbreaks resulted in 596 cases and 273 deaths.[2] In this latest EVD outbreak due to SUDV, the MOH rapidly acted to halt virus circulation in Kampala, as well as in all affected districts.[5, 13] Ebola Treatment Units were activated at Mubende and Fort Portal Regional Referral Hospitals on September 20, 2022, when the first case was reported. After the outbreak was confirmed, symptomatic contacts were evacuated directly to Ebola Treatment Units for testing. Activation of the Entebbe Regional Referral Hospital Ebola Treatment Unit in Wakiso District and Mulago National Referral Hospital Ebola Treatment Unit in Kampala District was on October 6, 2022, and October 15, 2022, respectively. Early on in the outbreak most cases represented health-care-associated rather than household transmission; cases due to burial, vertical or sexual transmission were rare.[13] The duration of the outbreak was 69 days, with a total of 164 and 77 confirmed cases and deaths recorded, respectively.[3] The mean age of cases was 28 years; among confirmed cases, 19 and 14 were health care providers and children <10 years of age, respectively; the highest case fatality ratio was observed among children <10 years of age (i.e., 75%) and adults between 40–49 years of age (i.e., 61.5%). The Ugandan MOH officially declared the end of the outbreak on January 11, 2023 (i.e., 60 days after the last infection event).

Following the report of the first case of SUDV, we developed an IBM-SUDV that modelled the burden and duration of the outbreak. Number of reported cases and deaths (164 and 77 reported *vs* 193 and 81 modelled), as well as a duration of the outbreak (16.5 reported *vs* 24 weeks modelled) was within the range of our Baseline Scenario. Our IBM-SUDV also showed that Delayed Outbreak and Out-of-Control Outbreak scenarios would have resulted in a substantially greater burden and duration of the outbreak, similar to the type of emergency experienced in 2014–2016 when an EVD outbreak resulted in 28,600 cases and 11,325 deaths across Guinea, Liberia and Sierra Leone. The results of our model highlighted that a rapid response to the EVD outbreak would be a crucial factor in Uganda’s ability for optimally effective control —which was what ultimately occured.[5, 13] The effective response was likely aided by: Uganda’s prior experience with responding to EVD outbreaks;[3] the development of a national disaster preparedness and management policy;[14] the establishment of a public health emergency operations centre and relevant task forces and other intra-government agency coordination bodies;[15] coordination of external partners; as well as infrastructures and resources build up in response to the COVID-19 pandemic.[7] Further modelling using the IBM-SUDV is currently being discussed with the MOH, including to estimate the potential effectiveness of changing various NPI parameters, assessing additional NPIs, as well as therapeutic options should these become available (e.g., monoclonal antibodies, vaccine).

## Data Availability

All data produced in the present work are contained in the manuscript.

## Acknowledgements

Funding for the work reported was provided by the US National Institute of Allergy and Infectious Disease/National Institutes of Health (NIAID/NIH), grants number U01AI51378 to the Center for Research in Emerging Infectious Diseases Coordinating Center (DB and RR) and grants number U01AI151799 to the Centre for Research in Emerging Infectious Diseases - East and Central Africa (CN, RB, NK).

## About the Author

Dr Bisanzio is a Senior Epidemiologist at RTI International. His primary research interests are the prevention, control, and surveillance of vector-borne, zoonotic, and emerging infectious diseases.

## 1 SUPPLEMENTARY METHODS

ABBREVIATIONS
|  |  |
| --- | --- |
| CDC | Center for Disease Control and Prevention |
| CrI | credible interval |
| CIESIN | Connectivity Lab and Center for International Earth Science Information Network |
| FW | frontline worker |
| GP | general population |
| GRASS | Geographic Resources Analysis Support System |
| HCW | healthcare worker |
| IBM | individual-based model |
| MAP | Malaria Atlas Project |
| NPI | nonpharmaceutical interventions |
| PPE | personal protection equipment |
| SD | standard deviation |
| SUDV | Sudan ebolavirus disease |
| UGA | Uganda |
| UNICEF | The United Nations Children's Fund |
| WHO | World Health Organization |

### 1.1 Model Overview

An individual-based model (IBM) was developed to (1) estimate the burden of cases and deaths, as well as (2) the duration of the unfolding 2022 Sudan ebolavirus (SUDV) disease outbreak in Uganda (UGA). Additionally, we evaluated the possible effect of non-pharmaceutical interventions (NPIs) on outbreak dynamics. The IBM-UGA structure was based on a contact network representing interactions of people at the local level (e.g., within households, schools, and local businesses) and at the regional level (e.g., movement between villages and cities). The resolution levels allow the model to account for contact heterogeneity among people while maintaining a manageable level of abstraction (and subsequently less computational complexity and faster run times).

The IBM contact network includes one node per person in the Uganda population, estimated to be 47.3 million people (World Bank, 2022). The links between nodes represent possible contacts between people and were created using publicly available and free geodata obtained by remote sensing and scientific literature. Geodata used by the model included population density maps produced by WorldPop (Lloyd et al., 2019) and Facebook (Facebook CIESIN, 2022) as well as accessibility maps produced by the Malaria Atlas Project (MAP) (Weiss et al., 2018).

**Figure 1.**
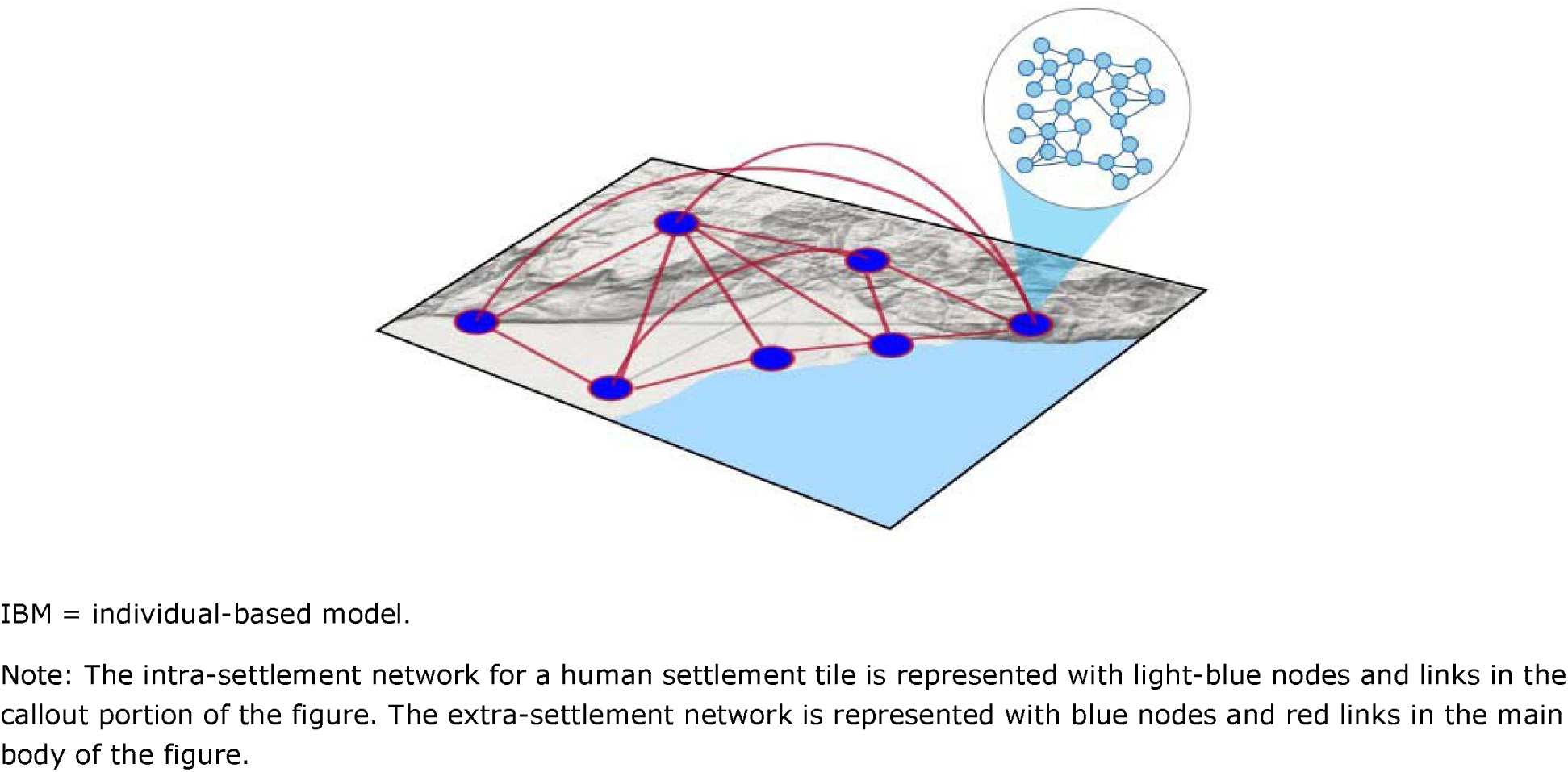
**Representation of the Spatial Network Used in the IBM to Represent Intra-settlement and Extra-settlement Contact**

The following provides a brief overview of the steps used to create the IBM:

Identified human settlements (cities and villages), including their geographical size (km2) and population, using density maps obtained from WorldPop (2022). To improve the model run time, the country was then divided into 10 km x 10 km tiles. The tiles represented the “settlement” units in the IBM. Villages typically fit within one tile, while a city may span many tiles. The density was adjusted to reflect the estimated population in 2022.

Created a dynamic network for each human settlement tile representing transmissible contacts of people on a per-week basis (intra-settlement network) and parameterized the network using information published in the scientific literature; this network represented short-range movement (Figure 1). The IBM accounted for changes in contact during an outbreak and included preintervention and postintervention periods to account for interventions adopted during an outbreak to reduce the spread of the disease.

Created links among people living in different human settlement tiles (extra- settlement network) using data from MAP’s accessibility map and WorldPop’s migration map and further merged with information from scientific literature; the links created in this process represent in-country, long-distance movement (Figure 1).

The human social network and the data used to characterize the network are described in more detail in Section 1.2 and Section 1.3.

SUDV transmission in the IBM was modeled using the classical SEIR compartmental model structure: Susceptible (S) ➔ Exposed (E) ➔ Infectious (I) ➔ Recovered (R) (Figure 2).

**Figure 2.**
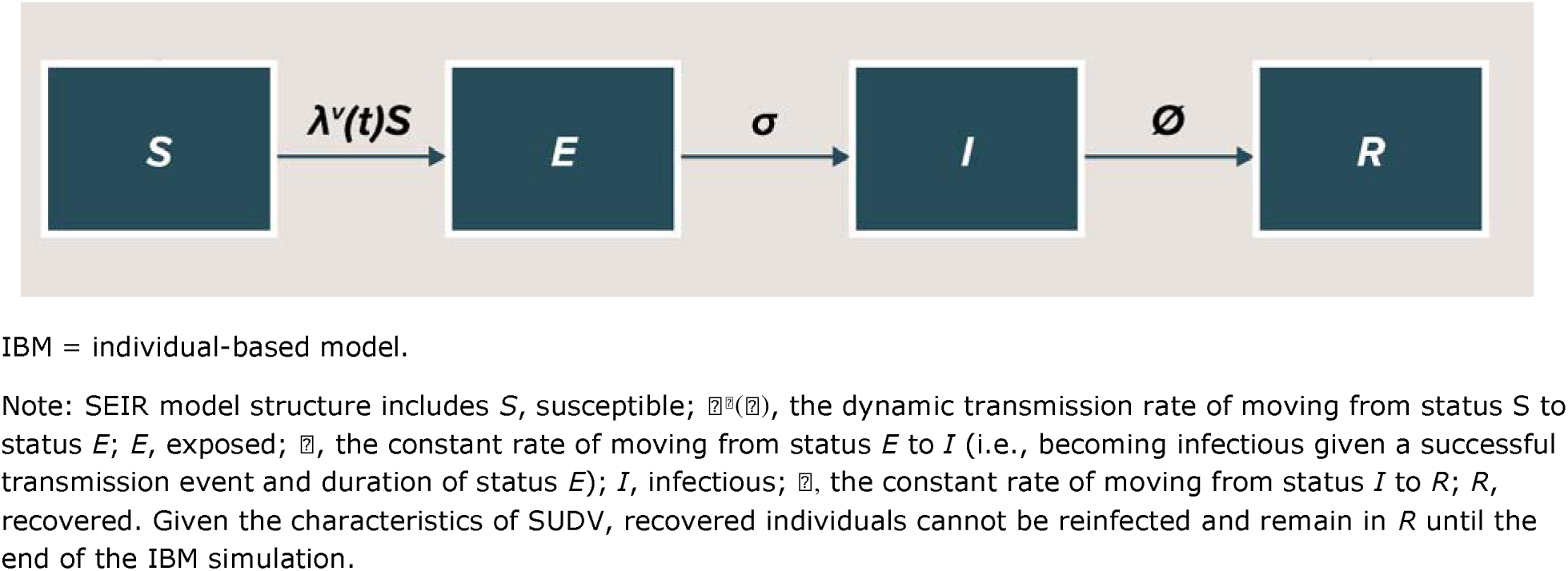
Model Structure for SUDV Dynamic Transmission IBM.

Infectious individuals were at risk of dying, and individuals who died remained infectious until they were buried. The transition from one status to another was a function of pathogen characteristics (e.g., probability of effective transmission per close contact, incubation period, infectious period, and fatality rate [see Table 3]) and interaction among individuals (only for S to I).

The following steps were taken to allow the IBM to account for SUDV transmission:

Estimated the number of treatment units (clinics and hospitals, not specific to SUDV) and healthcare workers (HCW) per human settlement tile, based on publicly available data from the World Health Organization (WHO) and published literature, to allow the IBM to represent human treatment-seeking behavior in the case of symptomatic Ebola virus disease (EVD).

Introduced the transmission of SUDV in the IBM using parameters from published literature.

Introduced the modification of human behavior (i.e., a dynamic social network) linked to SUDV infection.

Once the IBM was programmed with SUDV transmission, model outcomes were validated against available observed data.

Free, open-source software was used to program the IBM. The geodata was analyzed and stored using the free, open-source software called Geographic Resources Analysis Support System (GRASS) (GRASS Development Team, 2024). Data analysis was programmed using the R programming language (R Core Team, 2024). The IBM structure was programmed using the Julia programming language. Julia is a programming language suitable for IBMs because of its high speed, easy-to-use packages, and clear writing style (Bezanson et al., 2012). The IBM-UGA was run on a high-performance computer.

### 1.2. Human Social Network Data

#### 1.2.1 Population Data

Data on the Ugandan population were obtained from World Bank (World Bank, 2022) and disaggregated at into 10 km x 10 km tiles using data obtained from WorldPop (University of Southampton, 2020). WorldPop provided population estimates broken down by gender and age groupings (including 0–1 years and by 5-year ranges up to 80+ years) at a resolution of 100 meters. The estimates per each tile were adjusted to represent estimated population of 2022. The data obtained from WorldPop were used to estimate the size of the network for each human settlement.

#### 1.2.2 Human Settlements

To identify human settlements at high resolution, the population density map created by Facebook (Facebook CIESIN, 2022) was used. This map identified areas with buildings at a resolution of 30 meters, as updated to 2019. Population density map data were analyzed using spatial analysis techniques to identify the position of human settlements. To improve the model run time, the country was then divided into 10 km x 10 km tiles. The population in each tile was calculated by aggregating values from the density maps. The tiles represented the “settlement” units in the IBM—villages typically fit within one tile, while a city may span many tiles. A connectivity matrix among the tiles was created to represent the extra- settlement movement of individuals in Uganda.

#### 1.2.3 Travel Time Between Settlements

The movement of people between human settlement tiles was based on population size and travel times. Population sizes were calculated using WorldPop and Facebook data, as described previously. Travel times were calculated using the friction surface map created by MAP (Weiss et al., 2018). The friction surface map contains the time that a person spends passing through a map pixel based on estimated land-based travel speeds at a resolution of 1 km. Using the friction surface map estimation, we calculated the travel time between tiles.

#### 1.2.4 Demographic Data

The model accounted for several demographic characteristics that well represent the Ugandan population. Data on household size, percentage of children enrolled in schools, and the unemployment rate were obtained from the Global Data Lab (2016), the United Nations Children’s Fund (UNICEF) (2020), and the World Bank (2022). Additional population characteristics needed to define vaccination scenarios for preventive vaccination, such as the number of HCW, security forces personnel, and transportation workers, were also obtained from World Bank and WHO sources, as described in Section 1.3.2 .

### 1.3 IBM Population Network Structure

The IBM uses a dynamic human social network to describe interaction among individuals. The structure of the network is initially fixed, with all links between individuals assigned before the simulations begin. During each simulation run, the model becomes dynamic as EVD cases cause links to be “activated.” For example, infected individuals can acquire links as they travel to find hospitals, interact with HCWs, or interact with frontline workers (FWs) outside their fixed network. Thus, all simulations start with same initial network, but the network evolves differently during each simulated epidemic. The use of a fixed initial network reduces result noise and reflects the normal routine of a country during non- outbreak periods.

Given the SUDV transmission dynamics and the time lag of the surveillance to report cases, the model ran on a weekly time step. Weekly estimates also made it easier to compare IBM estimates with real data. The following sections provide details about the IBM human social network and its components.

#### 1.3.1 Representing the Human Social Network (Intra-settlement Network)

The IBM captured the high heterogeneity of the contact network among people, which is the key driver of disease spread in communities. The IBM network was built using the “scale-free” and “small-world” characteristics described for several social networks. A scale-free network has a high fraction of nodes (representing individuals) connected to a low number of other nodes and a few nodes connected to high number of other nodes. The nodes that have high connectivity are so-called “super spreaders.” The link distribution among nodes in the scale-free network followed a power-law distribution:

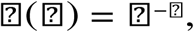

[where x is the number of links of a node, p(x) is the probability distribution, and the exponent α is the scaling factor. For human social networks, the power-law distributions have exponent α values ranging from 2 to 3 (Barrat et al., 2008). A network has a “small- world” characteristic when two nodes in the network can reach each other through a short sequence of connected nodes (called a “short path”) (Kleinberg, 2002).

Interactions among individuals occurred in specific locations, which may have a key role in the spread of disease agents. The locations in which people spend most of their time daily are households, workplaces, and schools (González et al., 2008; Wang et al., 2015).

However, interactions outside the routine locations (e.g., markets, restaurants, and theaters) are at the base of the small-world characteristic of human social networks. Age is another important factor that shapes the social network of an individual. People tend to have more interaction with individuals of the same age that they meet at school, workplaces, or in recreational locations (Prem et al., 2021). Thus, to be sure that the IBM accurately described the interaction among people, it included the distribution of interaction among individuals, interaction at locations, and the effect of the individuals’ ages.

After the population size and distribution among age groups within a settlement tile were estimated, a contact network was created using the following steps:

Households were created using the mean size of the family cluster. The number of people living together in each house was determined using a Poisson distribution with the mean equal to the mean number of household inhabitants in the Uganda by sub-regions as reported in the Uganda National Household Survey 2019/2020 (national mean household size was equal to 4.6 inhabitants) (Uganda Bureau of Statistics, 2021)).

The number of links (i.e., contacts) for each individual was calculated based on a power-law distribution with α = 2.5. Because the formulation of a power-law distribution requires indicating a minimum value for x, the minimum number of contacts for each individual was set equal to the mean household size in Uganda.

Once the number of links was determined for each individual, the distribution of these links to other individuals followed age-stratified contact matrices reported for Uganda in Prem, et al. (2021).

A location attribute was assigned to each link based on the settlement tile in which the contact occurred. The IBM also assigned one of four location types to each link: household, workplace, school, or other. The probability of assigning a link to a particular group was based on demographic characteristics of the Ugandan population.

Links defined by the power-law distribution represent contacts based on proximity rather than on close contact (e.g., touching). SUDV transmits through close contact, so the proportion of close contacts among all contacts (0.09) was estimated in preparation for SUDV transmission occurring (Olu et al., 2016; Wolfe et al., 2017).

#### 1.3.2 Representing the High-Risk Population

Within each settlement tile, the model accounted for the fraction of the population that are at high risk of infection during Ebola outbreaks due to their job activities. The IBM high-risk population included HCWs (e.g., doctors, nurses, midwives, dentists, and pharmacists) and non-health FWs (e.g., armed forces and transportation workers). The probability of infection for this population was affected by the probability of being in close contact with an infectious individual. As infectious individuals seek treatment in hospitals or other health facilities, the IBM’s treatment-seeking module creates links to connect infectious individuals with HCWs and FWs. These created links establish the exposure to the virus for the high-risk population. Linear regression was used to estimate the trend in the number of HCWs per profession to estimate figures for 2022, when not available.

The number of non-health FWs was estimated by type (i.e., armed forces, transportation workers, and other FWs) from published and public sources where possible (Table 1). No data were available for the number of other FWs, such as shop and market workers, clergy, contact tracers, and burial teams, so the model assumed a density of 3.5 per 1,000, which is between the density of HCWs (4.2 per 1,000) and transportation workers (2.5 per 1,000).

Next, to reflect that non-health FWs typically have many contacts in the community, these workers were assigned in the network from among individuals with the greatest number of contacts. Specifically, the IBM randomly selected individuals to be FWs from among those individuals in the 80th percentile for number of contacts. This selection was completed for each tile.

Table 1 reports the number of individuals in each high-risk population group.

**Table s1.**
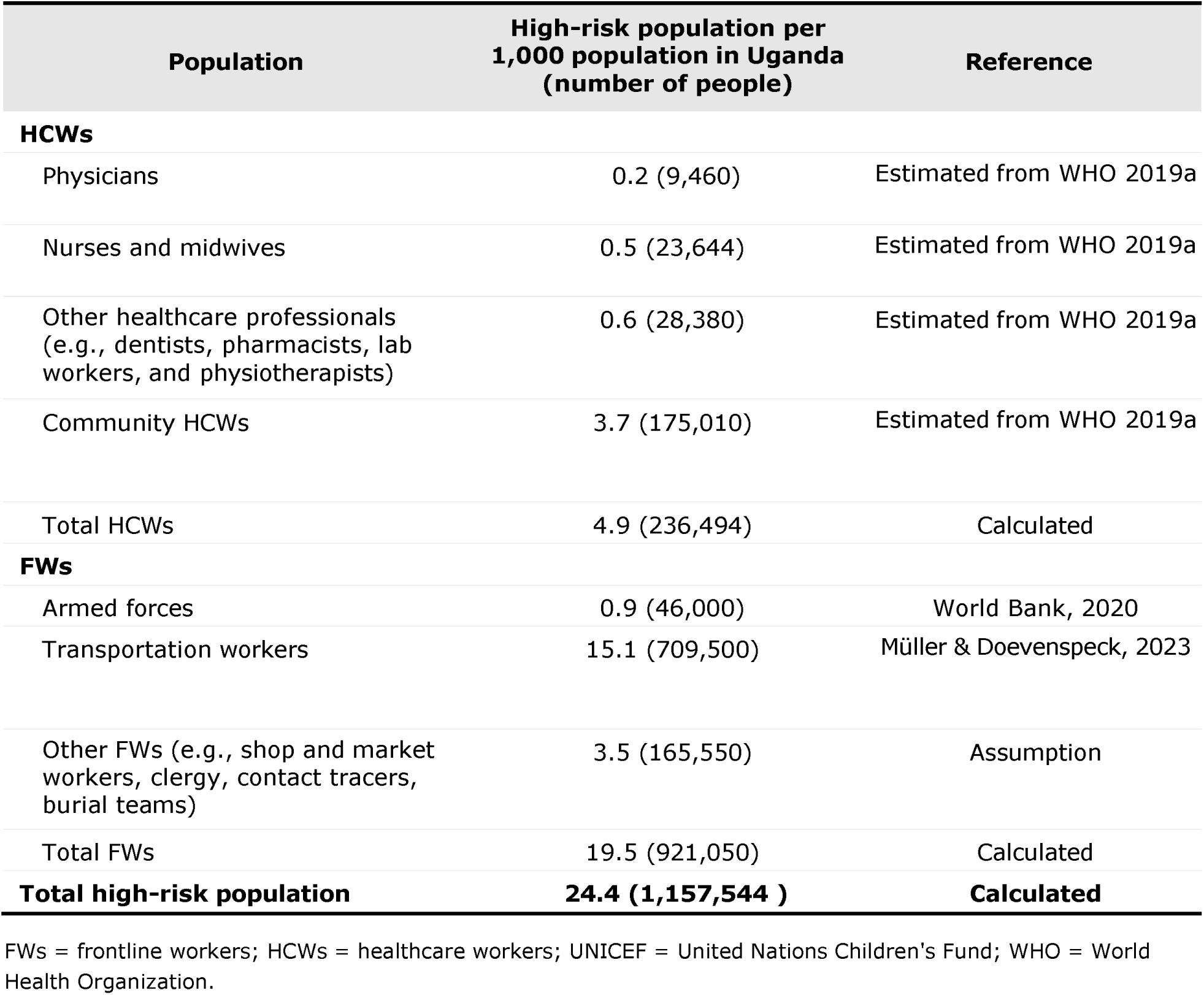
Uganda High-risk Population Estimated for the Year 2022.

#### 1.3.3 Representing Human Movement Among Settlements

The extra-settlement spread of SUDV in the IBM was captured using a weighted network that links settlement tiles (Figure 1). The weight of each link was determined by the estimated flux of people between tiles. Data about individual movement within Uganda were not available, so the IBM applied a gravity model that accounted for distance between pairs of settlements, travel times, and population sizes (Balcan et al., 2009; Kraemer et al., 2019). A gravity model is a modified law of gravitation that, in its simpler formulation (frictionless gravity model), considers the population size of two places and their distance apart to estimate the flow of people between them (Anderson, 2011). This approach is not country specific but has been used in the past in West Africa (Kraemer et al. 2019). Larger settlements were assumed to attract more people than smaller settlements, and settlements closer together were assumed to share more people than settlements further apart.

However, in settings where connections among places are not easy, the model can be adjusted by adding travel times (friction-based gravity model) (Anderson, 2011).

The distance and travel time between each pair of settlement tiles was calculated using the population density map (Facebook CIESIN, 2019) and the surface friction map (Weiss et al., 2018). The weight of each link was used to calculate the probability that Ebola cases will move between two settlement tiles, causing the outbreak to spread to new locations. The extra-settlement network was created using the following steps:

Created a distance matrix among settlement tiles with columns and rows equal to the number of settlement tiles. Each matrix cell contained the distance between two settlement tiles.

Created a travel time matrix among settlement tiles with columns and rows equal to the number of settlement tiles. Each matrix cell contained the travel time between two settlement tiles.

Built a gravity model merging population data, the distance matrix, and the travel time matrix following the methods described in Balcan et al. (2009) and Kraemer et al. (2019).

Recorded the results of the gravity model in a flux matrix with columns and rows equal to the number of settlement tiles. Each matrix cell contained the estimated flow between two settlement tiles. This matrix is not symmetric because the model uses tile population as an attraction factor. Thus, tiles with high population density have a greater inward flow than tiles with low population density.

Table 2 summarizes the parameters and data sources for the IBM intra-settlement and extra-settlement networks.

**Table s2.** IBM Population Network Parameters and Data Sources.

| Model input | Input | Reference |
| --- | --- | --- |
| Population | Estimated from the WorldPop (2020) | Lloyd et al., 2019 |
| Demographic data | World Bank Open Data, UNICEF (2019) | UNICEF, 2020 |
| Intrasettlement network |  |  |
| Number of contacts per individual | Estimated using a power-law distribution with $\alpha = 2.5$ | Barrat et al., 2008 |
| Contact matrix | Age-stratified, estimated | Prem et al., 2021 |
| Probability that a contact is a close contact | 0.09 | Estimated from Olu et al., 2016; Wolfe et al., 2017 |
| Extra-settlement network |  |  |
| Number of settlements in Uganda | Estimated from the Facebook density map | Facebook CIESIN, 2019 |
| Distance between settlements | Estimated from the Facebook density map | Facebook CIESIN, 2019 |
| Travel time between settlements | Estimated using MAP friction map | Weiss et al., 2018 |
| Extra-settlement flow of people | Estimated using the gravity model | Balcan et al., 2009; Kraemer et al., 2019 |

CIESIN = Connectivity Lab and Center for International Earth Science Information Network; IBM = individual-based model; MAP = Malaria Atlas Project; UNICEF = The United Nations Children's Fund.

### 1.4 Modeling SUDV Transmission

#### 1.4.1 Background

SUDV transmission depends on disease-specific characteristics, such as transmission probability per close contact, incubation period, and infectious period. Transmission is also affected by factors linked to population demographics and behavior (e.g., characteristics of the social network, treatment-seeking behavior, and burial practices), individual mobility, and healthcare system capacity. The IBM accounted for these factors to modulate the dynamic transmission of Ebola and affect the size of outbreaks. The model also included preintervention and postintervention periods to account for interventions adopted during an outbreak to reduce the spread of the disease, including ring vaccination, safe burials, contact tracing, extensive testing, HCW training, and availability of personal protection equipment (PPE). Section 1.4.2 provides information about the modeling of SUDV transmission, and Section 1.4.3 provides details about SUDV interventions included in the model.

#### 1.4.2 SUDV Transmission

In the model, each simulation starts with one infected individual selected at random. SUDV then transmits within the population as individuals interact. Because close contacts between individuals may vary in duration and proximity, only a fraction result in transmission. The probability of effective transmission per close contact is randomly selected for each simulation (one value for the entire simulation) between 0.45 and 0.55 (uniform distribution) (Table 3). Once infected, individuals with new cases of EVD may experience hospitalization or death.

Some individuals have infection-conferred immunity after surviving an Ebola infection during a previous outbreak. Since 2000, a total of 596 cases and 273 deaths have been reported in Uganda (CDC, 2020). Assuming 20% of cases were asymptomatic (estimated from Richardson et al., 2016; Mbala et al., 2017), the number of recovered cases since 2000 is 442. Accounting for deaths (CDC, 2020), an estimated 220 individuals are currently living with infection-conferred SUDV immunity in Uganda (Table 3). A summary of the parameters and data sources for Ebola transmission is provided in Table 3 .

**Table s3.** SUDV Transmission Parameters and Data Sources.

| Model input | Values/distribution | Reference |
| --- | --- | --- |
| Probability of effective transmission per close contact | 0.19–0.12<br>Uniform distribution | Estimated from Xia et al., 2015; Rivers et al., 2014 |
| Incubation period (days to symptom onset) | Mean: 12 days<br>SD: 4.3 days<br>Lognormal distribution | Estimated from Xia et al., 2015; Eichner et al., 2011 |
| Infectious period | 2–21 days<br>Uniform distribution | Xia et al., 2015; Malvy et al. 2019; Rivers et al., 2014 |
| Percentage of asymptomatic cases | 20% | Estimated from Richardson et al. 2016; Mbala et al., 2017 |
| Time from symptom onset to hospitalization | 5 days | Conrad et al., 2016 |
| Days in hospital | 10–15 days<br>Uniform distribution | Xia et al., 2015 |
| Case fatality rate by age | 39% | Estimated from WHO, 2022 |
| Time from symptom onset to death | 10 days | Legrand et al., 2007; WHO Ebola Response Team, 2015 ; Robert et al. 2019 |
| Days infectious before burial | 7 days | Estimated from Conrad et al., 2016; Potluri et. al 2020 |

SD = standard deviation; WHO = World Health Organization.

#### 1.4.3 SUDV Interventions

The model included NPIs as implemented during the last SUDV outbreaks in Uganda; a therapy or vaccine against SUDV disease does not exist. Interventions from the 2018-2020 outbreaks in the DRC were chosen as it was assumed that similar interventions would be used in Uganda, in accordance with current guidelines (WHO, 2021). Parameters for all interventions are reported in Table 4 .

NPI included the use of PPE for HCWs, contact tracing of infected individuals, safe burials, and increased treatment seeking due to increased awareness in the population (Table 4). These interventions were simulated using information about the strategies employed during the last EVD outbreaks in Uganda. To capture this heterogeneity, each settlement tile was randomly assigned a percentage between 20% and 70% (uniform distribution) of contacts of infected individuals successfully located by contact tracing teams (estimated from WHO, 2019b, and WHO, 2020). In the IBM, when a new case of EVD was identified within a settlement tile, linked contacts of the infected individual were randomly selected using this contact-traced percentage and designated as the contact-traced group. Individuals in this group represent those who were monitored for 21 days and hospitalized if showing any symptoms. Thus, individuals in the contact-traced group were not able to infect others.

Overall, NPIs were started in the model 1 week after the first reported case. Full implementation of NPIs in the model was reached within 40 weeks.

**Table s4.**
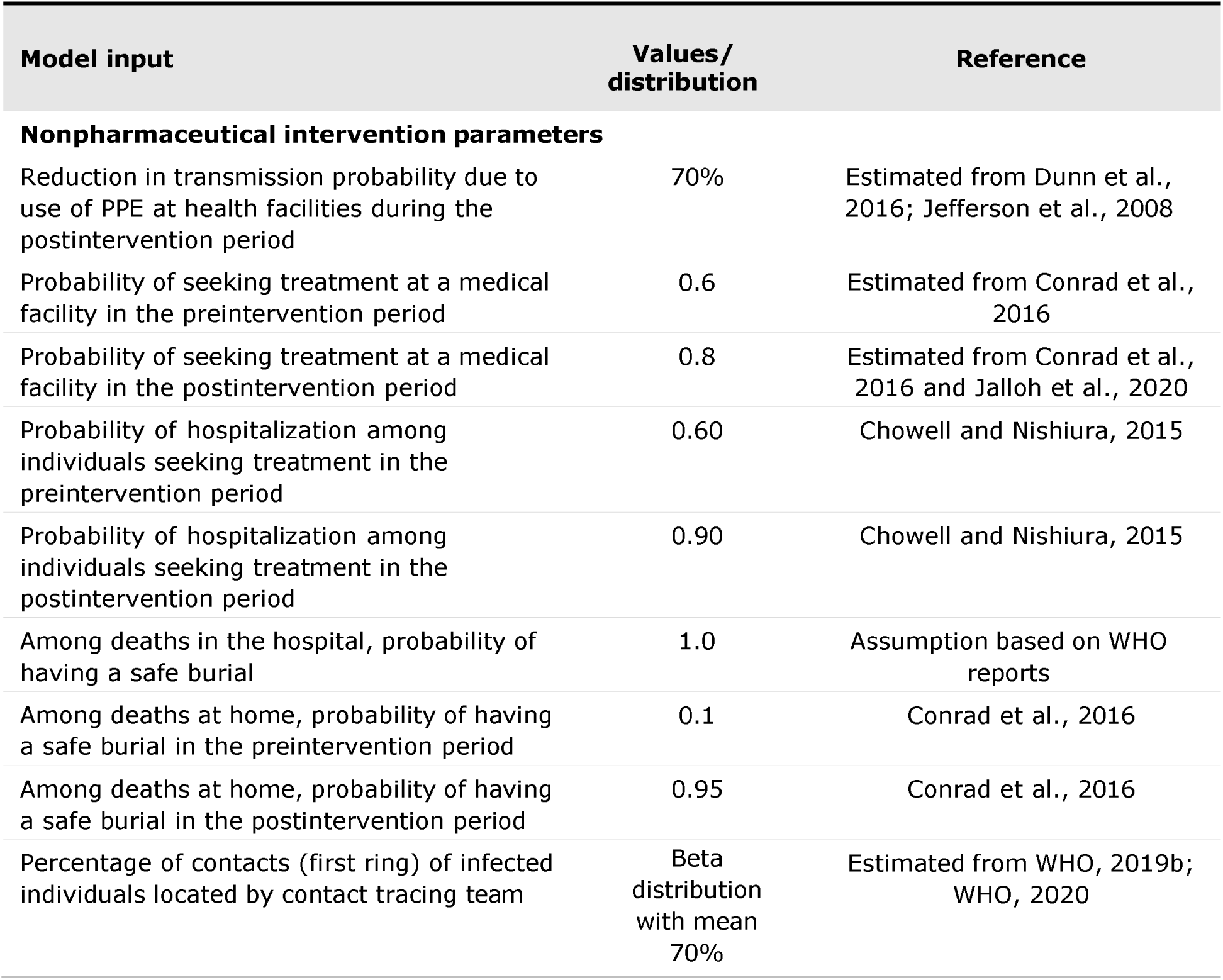
Parameters of Ebola Interventions.

### 1.5 IBM Assumptions

The model included several assumptions to populate variables for which data were not available, including the following:

As it is not possible to define urban and rural areas explicitly, settlements with over 10,000 inhabitants were assumed to be urban areas.

At least one healthcare worker was present in each settlement.

Hospitals and clinics with sufficient capacity to handle Ebola cases were located only in urban settlements. Cases requiring hospitalization were in contact with healthcare workers in the closest urban settlement.

Because the size of hospital crews vary, the model assumed the size of hospital crews followed a truncated Poisson distribution, with at least 2 people and a mean of 3 people (information obtained by personal communication with Médecins Sans Frontières personnel).

Armed forces were present in urban settlements, and personnel were assigned proportionally to the settlement’s population size.

The number of people who had contact with the body during a funeral followed a Poisson distribution with a mean of 10. This assumption was relevant only for traditional burials, as the model assumed transmission did not occur during safe burials.

### 1.6 Model Outcomes

The IBM estimated the median number of symptomatic cases and deaths as well as the median duration of the epidemic for each scenario analyzed. The epidemic duration was measured from the first case to the time at which zero cases remained.

Additionally, a 95% credible interval (CrI) was calculated for each outcome. CrIs are estimated in situations where the parameter is a random variable. CrIs then represent the interval within which an unobserved parameter value falls with a given probability. In comparison, confidence intervals are estimated in situations with known, observed data, where intervals can be precisely calculated. For the IBM, 95% CrIs were calculated using the adjusted bootstrap percentile approach (Davison and Hinkley, 1997) based on 10,000 re- samplings of the simulation results. This bootstrapping allows better estimates of the 95% CrIs because it calculates the CrIs from an estimated hypothetical distribution of the results.

Finally, the reduction in each outcome (and the 95% CrI of the reduction) was calculated for the comparison between several of the scenarios. The reductions and their 95% CrIs were calculated using the bootstrapping method described above (Davison and Hinkley, 1997). Because of this, the reported reductions are not equivalent to the corresponding reductions in the median outcomes.

